# Impact of COVID-19 non-pharmaceutical interventions on pneumococcal carriage prevalence and density in Vietnam

**DOI:** 10.1101/2022.05.05.22274646

**Authors:** Monica Larissa Nation, Sam Manna, Hau Phuc Tran, Cattram Duong Nguyen, Le Thi Tuong Vy, Doan Y Uyen, Tran Linh Phuong, Vo Thi Trang Dai, Belinda Daniela Ortika, Ashleigh Christina Wee-Hee, Jemima Beissbarth, Jason Hinds, Kathryn Bright, Heidi Smith-Vaughan, Thuong Vu Nguyen, Kim Mulholland, Beth Temple, Catherine Satzke

## Abstract

Non-pharmaceutical interventions (NPIs) implemented to contain SARS-CoV-2 have decreased invasive pneumococcal disease. We undertook an observational study to evaluate the impact of NPIs on pneumococcal carriage and density, drivers of transmission and disease, during the COVID-19 pandemic in Ho Chi Minh City, Vietnam. While NPIs did not significantly impact pneumococcal carriage, mean capsular pneumococcal density decreased by up to 91.5% (1.07 log_10_genome equivalents/mL, 95% Confidence Interval: 0.74-1.41) after NPI introduction compared with the pre-COVID-19 period. As higher pneumococcal density is a risk factor for disease, the observed decline provides a plausible mechanism for the reductions in invasive pneumococcal disease.

## INTRODUCTION

*Streptococcus pneumoniae* is a major cause of disease including pneumonia, meningitis, and sepsis. The bacterium commonly colonises the nasopharynx and carriage is required for pneumococcal transmission and disease.[1]

During the COVID-19 pandemic, implementation of non-pharmaceutical interventions (NPIs) such as mask wearing, stay-at-home orders, and physical distancing have been critical for the containment of SARS-CoV-2. Introduction of NPIs have been associated with declines in other respiratory pathogens and infectious diseases, including influenza, respiratory syncytial virus, pneumonia, and invasive pneumococcal disease (IPD).[2–4] Only two studies have examined the impact of NPIs on pneumococcal carriage prevalence.[4,5] No substantive differences in carriage prevalence were observed in children pre- and post-implementation of NPIs in Belgium and Israel, but a decline in the prevalence of respiratory viruses was temporally associated with a decline in IPD in the Israel study.[4] Respiratory viruses increase pneumococcal carriage density, which contributes to transmission and disease.[6,7] Therefore, we hypothesized reductions in IPD associated with NPIs may be due to reductions in carriage density. Here, we aim to assess the impact of NPIs during the COVID-19 pandemic on pneumococcal carriage and density in children in Vietnam.

## METHODS

Nasopharyngeal samples were collected from children aged 24-months between 25 December 2018 and 18 June 2020 as part of a pneumococcal conjugate vaccine (PCV) trial in Ho Chi Minh City, Vietnam.[8] All children with a 24-month sample were eligible for inclusion in this study. Swabs were collected using WHO guidelines and pneumococcal carriage and density determined by quantitative PCR (qPCR) targeting the *lytA* gene and DNA microarray.[8] Three time periods were defined based on key events and the implementation of NPIs in Vietnam (refer to supplementary material): a pre-COVID-19 period (25 December 2018 - 31 December 2019), NPI period 1 (NPI-1, 03 February - 31 March 2020), and NPI period 2 (NPI-2, 23 April - 18 June 2020).

We determined the prevalence and density of overall, capsular, and non-encapsulated pneumococcal carriage. Participant characteristics were assessed using the chi-squared test for comparisons of proportions, or quantile regression with bootstrapped standard errors for comparisons of medians. Potential confounders were assessed using a directed acyclic graph, with no confounders identified (Figure S1). Density data were log_10_ transformed and are expressed as log_10_genome equivalents per mL. Pneumococcal carriage and density in each NPI period were compared with the pre-COVID-19 period using unadjusted logistic and linear regression. In a secondary analysis, the direct effect (i.e. not via intermediate variables) of the NPI period on pneumococcal carriage and density were evaluated by including district and season in the models. Analyses were repeated in unvaccinated children alone to confirm any differences were independent of the trial intervention. Statistical analyses were conducted using Stata v16.0 (StataCorp LLC).

## RESULTS

Over the three time periods, 2106 nasopharyngeal samples were collected (n=1537 pre-COVID-19, n=307 in NPI-1, and n=262 in NPI-2; Figure S2). Participant characteristics were similar across the time periods for sex, current breastfeeding, number of PCV doses received, and the type of PCV received (Table S1).

Of the 2106 children assessed, 485 (23.0%) carried pneumococci. There was no difference in overall pneumococcal carriage in NPI-1 compared with pre-COVID-19 (Odds Ratio (OR): 0.96 (95% Confidence Interval (95%CI): 0.71-1.28). There was some evidence of a reduction in overall carriage in NPI-2 compared with pre-COVID-19 (OR: 0.73 (95%CI: 0.52-1.01; p-value=0.060). Neither NPI period was associated with changes in capsular or non-encapsulated carriage (Table 1).

**Table 1.**
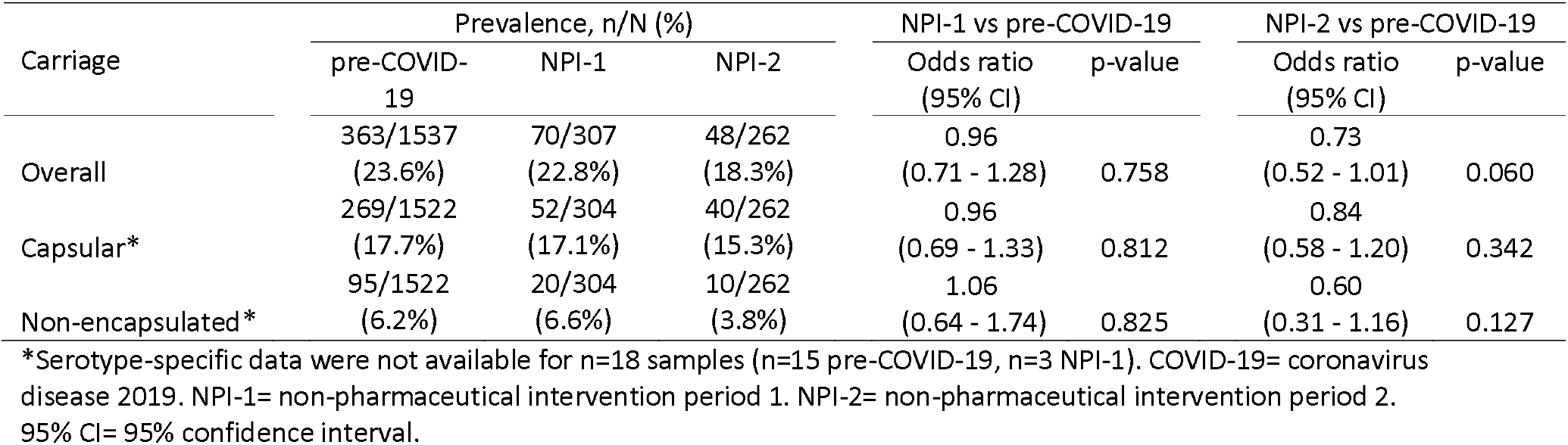
Pneumococcal carriage prevalence and odds ratios in periods with non-pharmaceutical interventions (NPIs) compared with the pre-COVID-19 period.

Overall and capsular pneumococcal carriage density decreased from pre-COVID-19 to NPI-1, with a further reduction observed in NPI-2 (Figure 1, Table 2). The decrease from pre-COVID-19 to NPI-2 was 85.5% (i.e. 0.84 log_10_ genome equivalents/mL, 95%CI: 0.55-1.13), and 91.5% (i.e. 1.07 log_10_ genome equivalents/mL, 95%CI: 0.74-1.41) for overall and capsular pneumococcal density, respectively. Reductions in overall pneumococcal density were driven by decreases in capsular pneumococcal density, with no corresponding reduction in non-encapsulated pneumococcal density. Similar patterns were observed in the secondary analysis adjusting for intermediate variables (Table S2), and in the analyses restricted to the unvaccinated group alone (Table S3).

**Table 2.** Pneumococcal carriage density means and difference in means in periods with non-pharmaceutical interventions (NPIs) compared with the pre-COVID-19 period.

| Density† | N | Mean (SD) |  |  | NPI-1 vs pre-COVID-19 |  | NPI-2 vs pre-COVID-19 |  |
| --- | --- | --- | --- | --- | --- | --- | --- | --- |
|  |  | pre-COVID-19 | NPI-1 | NPI-2 | Difference in means (95% CI) | p-value | Difference in means (95% CI) | p-value |
| Overall | 481 | 6.07<br>(0.99) | 5.63<br>(0.90) | 5.23<br>(0.92) | -0.44<br>(-0.69 - -0.19) | 0.001 | -0.84<br>(-1.13 - -0.55) | <0.001 |
| Capsular* | 361 | 6.18<br>(1.01) | 5.65<br>(0.95) | 5.11<br>(0.95) | -0.53<br>(-0.83 - -0.23) | 0.001 | -1.07<br>(-1.41 - -0.74) | <0.001 |
| Non-encapsulated* | 125 | 5.90<br>(0.74) | 5.60<br>(0.63) | 5.69<br>(0.52) | -0.29<br>(-0.64 - 0.05) | 0.098 | -0.21<br>(-0.68 - 0.26) | 0.381 |

†Assessed in pneumococcal carriers only and reported in log<sub>10</sub> genome equivalents per mL. \*Serotype-specific data were not available for n=18 samples (n=15 pre-COVID-19, n=3 NPI-1). NPI-1= non-pharmaceutical intervention period 1. NPI-2= non-pharmaceutical intervention period 2. COVID-19= coronavirus disease 2019. SD= standard deviation. 95% CI= 95% confidence interval.

**Figure 1.**
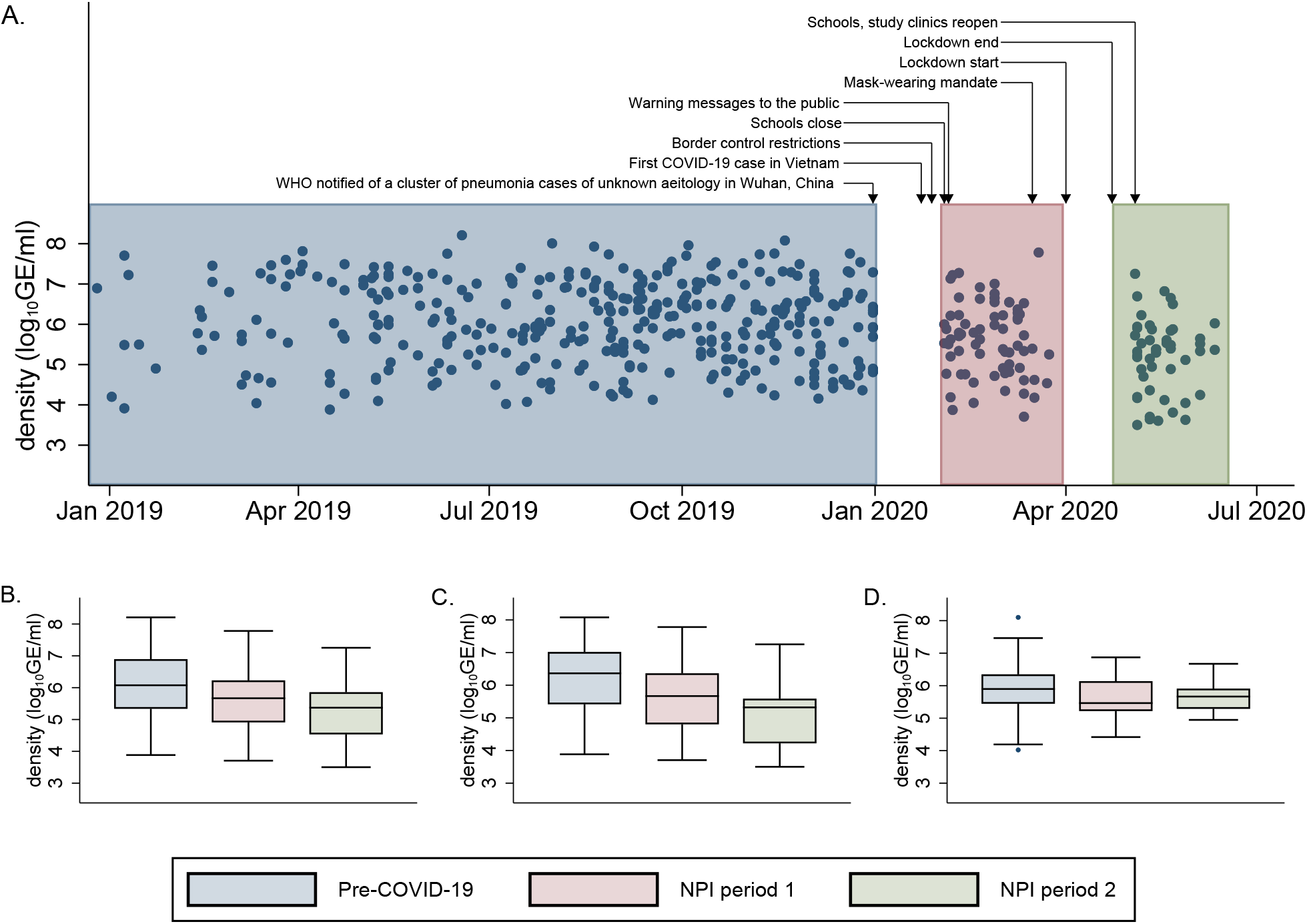
Nasopharyngeal carriage density (log_10_ genome equivalents per mL) among pneumococcal carriers. A) Overall pneumococcal density during 2019-2020 with key events and non-pharmaceutical interventions (NPIs) indicated. Box plots of B) overall, C) capsular, and D) non-encapsulated pneumococcal density. Boxes depict interquartile range (IQR) with a central line for the median. Data points further than the 25^th^/75^th^ percentile plus 1.5 times the IQR are plotted as individual points. COVID-19= coronavirus disease 2019.

## DISCUSSION

Following the implementation of NPIs for the containment of SARS-CoV-2, a decline in disease caused by other respiratory pathogens including pneumococci has been observed in multiple countries. The decline in IPD has been attributed to reductions in pneumococcal transmission.[3] We found some evidence of a 27% reduction in carriage, although this wasn’t observed in unvaccinated participants alone. Studies in Belgium and Israel found no substantive reductions in prevalence pre- and post-NPIs, although in both our study and the Israel study pneumococcal prevalence was lower in one of the two time periods assessed.[4,5] Importantly, we observed a substantial decline in overall and capsular pneumococcal carriage density after the implementation of NPIs. In contrast, the Israel study found no difference in pneumococcal density pre- and post-NPI implementation.[4] These differences may be partially explained by the use of the more sensitive quantitative PCR method in our study. It is unclear why there was no reduction in non-encapsulated density. Interestingly, we have previously found that co-infection of infant mice with a murine analogue of RSV increased the density of capsulated pneumococci, but this increase was not observed in non-encapsulated pneumococci.[9]

A major strength of this study is the use of carriage data from healthy children. Most studies investigating the impact of NPIs on non-SARS-CoV-2 pathogens rely on disease data, which can be influenced by behavioral changes such as reluctance to seek medical treatment for fear of contracting COVID-19. One limitation is that other temporal changes may have occurred during the study that were not accounted for.

The implementation of NPIs has been associated with suppression of circulating respiratory viruses.[4] Such viruses are known to increase pneumococcal carriage density,[6,7] which is a risk factor for disease.[7,10] The decline we observed in pneumococcal density associated with NPI implementation therefore provides a plausible mechanism for the reductions in IPD that have been observed elsewhere in the absence of a substantive reduction in carriage prevalence. Further research examining the links between pneumococcal carriage, density, and respiratory viruses is needed to fully elucidate the impact NPIs have on IPD.

## Data Availability

Deidentified data are available upon reasonable request. Requests must be compliant with the vaccine trial ethical approvals. Requests should be directed to Professor Kim Mulholland (https://orcid.org/0000-0001-7947-680X).

## Ethics statements

### Patient consent for publication

All participants gave consent for results to be published in scientific journals as part of the Participant Information and Consent form completed during enrollment into the vaccine trial.

## Ethics approval

Ethical approval was obtained for the vaccine trial from the Human Research Ethics Committee of the Royal Children’s Hospital Melbourne, the Institutional Review Board at the Pasteur Institute of Ho Chi Minh City, and the Vietnam Ministry of Health Ethical Review Committee for Biomedical Research. The vaccine trial is registered at clinicaltrials.gov (NCT03098628).

## Acknowledgments

We thank the study participants and their families, and the study staff. We acknowledge the contributions that Phan Trong Lan, Sophie La Vincente, Helen Thomson, Anne Balloch, Paul Licciardi, and Thi Que Huong Vu (deceased) made to the vaccine trial.

## Contributors

CS and MLN conceived this study with input from BT and SM. BDO and ACW conducted the laboratory analyses with oversight from CS. JH contributed to the interpretation of microarray data. MLN conducted the data analyses with input from CS, BT, and CDN. MLN, CS, and SM prepared the original manuscript. LTTV, TLP, and DY coordinated the trial sites, with oversight from HPT. KM and TVN are Principal Investigators of the vaccine trial, with contributions from BT, HPT, KB, VTTD, CS, HS-V, DYU, JB, and CDN. All authors reviewed and approved the final manuscript.

