## Supplementary Information for "Impact of COVID-19 non-pharmaceutical interventions on pneumococcal carriage prevalence and density in Vietnam"

**Additional methodological details**

Study participants

Nasopharyngeal samples were collected from children enrolled in a trial investigating reduced-dose schedules of pneumococcal conjugate vaccines.[1] In the trial, children were enrolled at 2 months of age and randomised to one of five groups and followed up to 24 months of age. Participants received PCV10 at 12 months (0+1) or at 2 and 12 months (1+1), or PCV13 at 12 months (0+1) or at 2 and 12 months (1+1), or a single dose of PCV10 at 24 months (control group). Nasopharyngeal sample collection for the 24-month timepoint occurred before and after the emergence of SARS-CoV-2, enabling the impact of non-pharmaceutical interventions (NPI) on pneumococcal carriage prevalence and density to be evaluated.

NPI periods

NPI periods were defined by key events and the implementation of NPIs in Vietnam.[2] The pre-COVID-19 period was defined from the start of the 24-month sample collection until the World Health Organization (WHO) was notified of a cluster of cases in Wuhan, China (25 December 2018 - 31 December 2019). The first NPI period included school closures, warning messages to the public, restrictions on non-essential businesses, and a mask wearing mandate (NPI-1; 03 February 2020 - 31 March 2020). A three-week lockdown was implemented in Ho Chi Minh City from 01 April to 22 April 2020, during which time study clinics were closed and no samples were collected. The second NPI period started after the lockdown and concluded at the end of sample collection (NPI-2; 23 April 2020 - 18 June 2020). Samples collected between 01 January 2020 and 02 February 2020 were excluded from analyses as preventative measures (such as mask-wearing, hand hygiene and self-isolation) may have been implemented before such policies were officially introduced.

Season

Ho Chi Minh City has a Tropical Savannah climate [3] with two seasons. The dry season is from November to April, and the rainy season is from May to October.

Serotyping interpretation and carriage outcomes

Molecular serotyping was performed using Senti-SPv1.5 DNA microarrays (BUGS

Bioscience), with analysis using a custom web-based software.[4] Serotype 11F-like was reported as 11A, and serotypes 15B and 15C were reported as 15B/C.[5,6] Overall pneumococcal carriage was defined as carriage of any pneumococci. Capsular carriage included carriage of any serotype excluding non-encapsulated pneumococci. Non-encapsulated carriage included carriage of previously described non-encapsulated pneumococci including NT2, NT3b, NT4a, NT4b.[7] Any sample containing pneumococci was considered positive. For example, a sample containing both capsular and non-encapsulated serotypes was considered positive for overall, capsular, and non-encapsulated carriage. Serotype-specific density was calculated by multiplying the overall pneumococcal load (as determined by qPCR) by the corresponding relative abundance of the serotype (as determined by microarray).

Directed acyclic graph (DAG)

A DAG was constructed to evaluate the relationships between the exposure (NPI period), outcomes (pneumococcal carriage and density), and other key factors. Arrows are used to indicate the direction of the association. Green lines indicate causal pathways, and red lines indicate biasing pathways (none identified). Measured variables are shown in dark grey and unmeasured variables are shown in light grey. No potential confounders were identified for the primary analysis, and therefore no covariates were included for adjustment. As the DAG indicates there are some intermediate variables on the causal pathway between exposure and outcome (i.e. some of the effect of NPI period on pneumococci could be mediated by district of residence and season), we chose to conduct a secondary analysis evaluating the direct effect of NPI period on pneumococcal carriage and density. In the secondary analysis, district of residence and season were included in the models for adjustment.

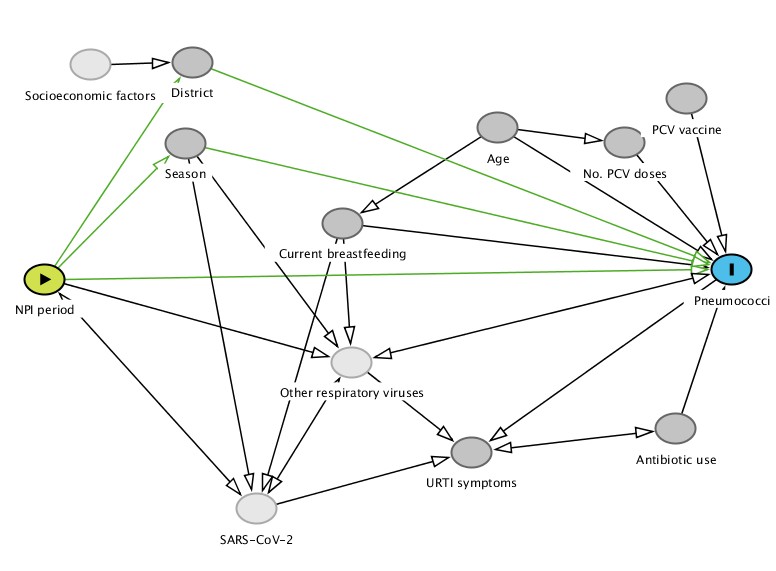

**Figure S1.** Directed acyclic graph depicting the relationships between the exposure (NPI period), outcomes (pneumococcal carriage and density), and other key factors. Green lines indicate causal pathways, and red lines indicate biasing pathways (none identified). Measured variables are shown in dark grey and unmeasured variables are shown in light grey.

**Additional results**

Participant flow and characteristics

Of the 2501 participants enrolled in the trial, 318 withdrew and do not contribute data to this analysis. Between 25 December 2018 and 18 June 2020, 2183 nasopharyngeal samples were collected from children aged approximately 24 months old. Seventy-seven samples were collected between 01 January and 02 February 2020 and were excluded from analysis. Across time periods participant characteristics were similar for sex, current breastfeeding, number of PCV doses received, and the type of PCV received. However, children were older, reported fewer respiratory symptoms, and used fewer antibiotics in NPI period 2 compared with the pre-COVID-19 period or NPI period 1 (Table S1). Samples were collected across both seasons in the pre-COVID-19 period, whilst all samples in NPI period 1 were collected in the dry season and all samples in NPI period 2 were collected in the rainy season. There were differences in the proportion of participants from each district as recruitment commenced at different times in the districts.

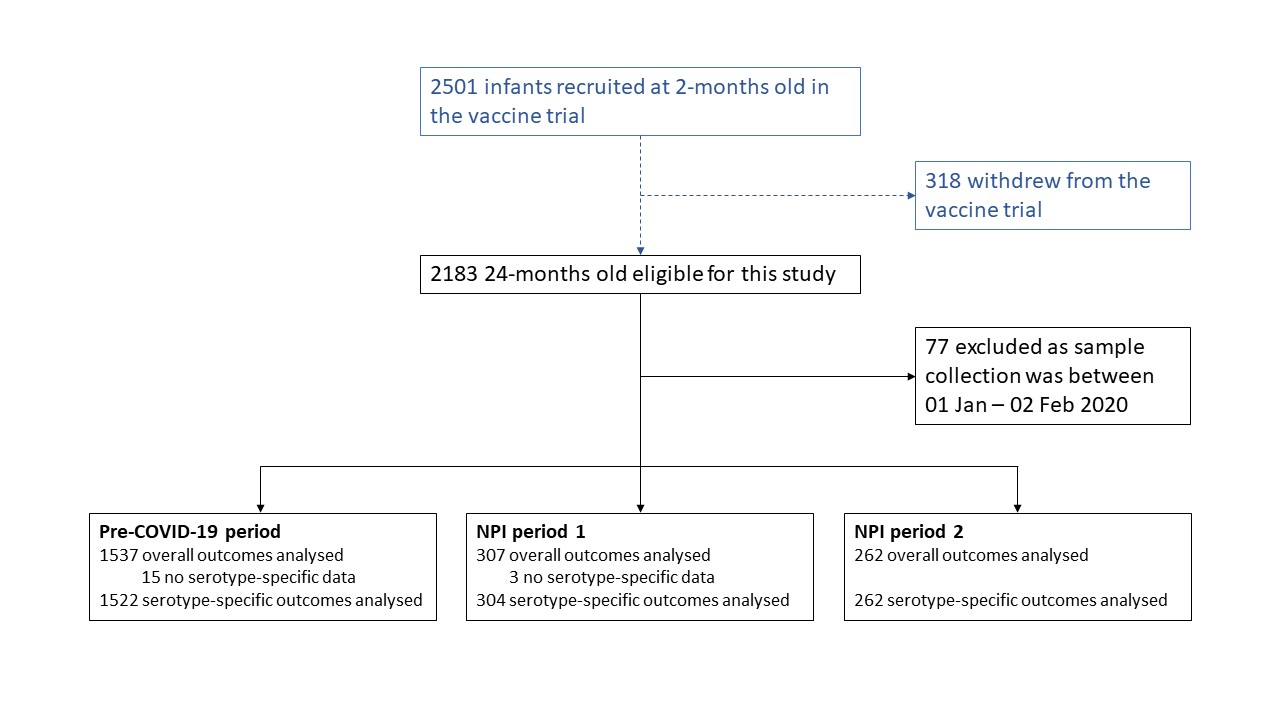

**Figure S2.** Participant flow. Participants were excluded from all analyses if samples were collected between 01 January - 02 February 2020 as preventative measures may have been implemented before the non-pharmaceutical interventions (NPI) were officially introduced (n=77), or were excluded from serotype-specific analyses if serotype-specific data was not available (n=18).

**Table S1.** Participant characteristics at the 24-month visit

|  | Pre-COVID-19 period  n=1537 | NPI period 1  n=307 | NPI period 2  n=262 | p-value* |
| --- | --- | --- | --- | --- |
| Age in months [median(range)] | 24.1 (24.0-28.5) | 24.1 (24.0-28.7) | 25.0 (24.0-30.8) | <0.001 |
| Sex, female | 740 (48.1%) | 148 (48.2%) | 131 (50.0%) | 0.860 |
| District |  |  |  | <0.001 |
| 4 | 663 (43.1%) | 88 (28.7%) | 74 (28.2%) |  |
| 7 | 381 (24.8%) | 104 (33.9%) | 68 (26.0%) |  |
| 8 | 493 (32.1%) | 115 (37.5%) | 120 (45.8%) |  |
| Season† |  |  |  | <0.001 |
| Dry | 513 (33.4%) | 307 (100%) | 0 (0%) |  |
| Rainy | 1024 (66.6%) | 0 (0%) | 262 (100%) |  |
| Current breastfeeding | 120 (7.8%) | 19 (6.2%) | 14 (5.3%) | 0.268 |
| Current URTI symptoms | 250 (16.3%) | 14 (4.6%) | 7 (2.7%) | <0.001 |
| Antibiotic use in past 2 weeks | 175 (11.4%) | 8 (2.6%) | 2 (0.8%) | <0.001 |
| Current antibiotic use | 62 (4.0%) | 5 (1.6%) | 0 (0%) | 0.001 |
| Doses of PCV received§ |  |  |  | 0.820 |
| 0 | 540 (35.1%) | 110 (35.8%) | 87 (33.2%) |  |
| 1 | 498 (32.4%) | 91 (29.6%) | 89 (34.0%) |  |
| 2 | 499 (32.5%) | 106 (34.5%) | 86 (32.8%) |  |
| If PCV received, PCV10 | 486 (48.7%) | 101 (51.3%) | 88 (50.3%) | 0.780 |

Data are n (%) unless specified. URTI= upper respiratory tract infection. PCV= pneumococcal conjugate vaccine. PCV10= ten-valent PCV.

*p-values are comparisons across all time periods based on chi-squared test (for comparisons of proportions), or quantile regression with bootstrapped standard errors (for comparisons of medians). †Dry season: November-April, rainy season: May-October. §Participants received either PCV10 or PCV13.

**Table S2.** Adjusted coefficients and 95% confidence intervals for pneumococcal carriage and density in periods with non-pharmaceutical interventions (NPIs) compared with the pre-COVID-19 period. Coefficients are odds ratios for pneumococcal carriage and difference in means for density.

|  | Adjusted* | | | | |
| --- | --- | --- | --- | --- | --- |
| Outcome | N | NPI period 1  Coefficient (95% CI) | p-value | NPI period 2  Coefficient (95% CI) | p-value |
| Carriage |  |  |  |  |  |
| Overall | 2106 | 0.79 (0.56 - 1.10) | 0.159 | 0.76 (0.54 - 1.08) | 0.122 |
| Capsular§ | 2088 | 0.78 (0.53 - 1.13) | 0.183 | 0.88 (0.60 - 1.28) | 0.497 |
| Non-encapsulated§ | 2088 | 0.99 (0.55 - 1.76) | 0.963 | 0.62 (0.31 - 1.23) | 0.173 |
| Density† |  |  |  |  |  |
| Overall | 481 | -0.35 (-0.64 - -0.06) | 0.017 | -0.85 (-1.16 - -0.55) | <0.001 |
| Capsular§ | 361 | -0.39 (-0.73 - -0.05) | 0.025 | -1.11 (-1.46 - -0.76) | <0.001 |
| Non-encapsulated§ | 125 | -0.31 (-0.72 - 0.10) | 0.133 | -0.21 (-0.69 - 0.28) | 0.394 |

*Adjusted for district of residence and season of swab collection to evaluate the direct effect of the NPI period on pneumococci.

§Serotype-specific data were not available for n=18 samples (n=15 pre-COVID-19, n=3 NPI-1).

†Assessed in pneumococcal carriers only and reported in log_10_ genome equivalents per mL.

95% CI= 95% confidence interval.

|  | Unadjusted | | | | | Adjusted* | | | | |
| --- | --- | --- | --- | --- | --- | --- | --- | --- | --- | --- |
| Outcome | N | NPI period 1  Coefficient (95% CI) | p-value | NPI period 2  Coefficient (95% CI) | p-value | N | NPI period 1  Coefficient (95% CI) | p-value | NPI period 2  Coefficient (95% CI) | p-value |
| Carriage  – unvaccinated only |  |  |  |  |  |  |  |  |  |  |
| Overall | 737 | 1.32 (0.84 - 2.08) | 0.223 | 1.11 (0.66 - 1.86) | 0.689 | 737 | 1.18 (0.69 - 2.00) | 0.550 | 1.14 (0.66 - 1.95) | 0.641 |
| Capsular§ | 730 | 1.31 (0.80 - 2.15) | 0.287 | 1.38 (0.81 - 2.37) | 0.235 | 730 | 1.10 (0.62 - 1.97) | 0.738 | 1.43 (0.81 - 2.51) | 0.219 |
| Non-encapsulated§ | 730 | 1.13 (0.48 - 2.63) | 0.783 | 0.38 (0.09 - 1.63) | 0.193 | 730 | 1.05 (0.39 - 2.82) | 0.923 | 0.40 (0.09 - 1.74) | 0.221 |
| Density†  – unvaccinated only |  |  |  |  |  |  |  |  |  |  |
| Overall | 188 | -0.39 (-0.78 - -0.00) | 0.048 | -0.82 (-1.27 - -0.37) | <0.001 | 188 | -0.27 (-0.73 - 0.19) | 0.254 | -0.86 (-1.34 - -0.38) | <0.001 |
| Capsular§ | 146 | -0.39 (-0.86 - 0.07) | 0.099 | -0.98 (-1.48 - -0.48) | <0.001 | 146 | -0.22 (-0.77 - 0.33) | 0.433 | -1.06 (-1.60 - -0.52) | <0.001 |
| Non-encapsulated§ | 40 | -0.15 (-0.66 - 0.36) | 0.557 | 0.02 (-0.87 - 0.91) | 0.966 | 40 | 0.11 (-0.49 - 0.71) | 0.712 | -0.10 (-0.98 - 0.78) | 0.823 |

**Table S3.** Unadjusted and adjusted coefficients and 95% confidence intervals for pneumococcal carriage and density in unvaccinated participants in periods with non-pharmaceutical interventions (NPIs) compared with the pre-COVID-19 period. Coefficients are odds ratios for pneumococcal carriage and difference in means for density.

95% CI= 95% confidence interval.

*Adjusted for district of residence and season of swab collection to evaluate the direct effect of the NPI period on pneumococci.

§Serotype-specific data were not available for n=18 samples (n=5 pre-COVID-19, n=2 NPI-1).

†Assessed in pneumococcal carriers only and reported in log_10_ genome equivalents per mL.
